# Safety and Immunogenicity of a Recombinant Protein RBD Fusion Heterodimer Vaccine against SARS-CoV-2: preliminary results of a phase 1-2a dose-escalating, randomized, double-blind clinical trial

**DOI:** 10.1101/2022.08.09.22278560

**Authors:** Lorna Leal, Judit Pich, Laura Ferrer, Jocelyn Nava, Ruth Martí-Lluch, Ignasi Esteban, Edwards Pradenas, Dàlia Raïch-Regué, Antoni Prenafeta, Karla Escobar, Carmen Pastor, Marc Ribas-Aulinas, Benjamin Trinitè, Jordana Muñoz-Basagoiti, Gemma Domenech, Bonaventura Clotet, Júlia Corominas, Aida Corpes-Comes, Carme Garriga, Antonio Barreiro, Nuria Izquierdo-Useros, Joan Albert Arnaiz, Alex Soriano, José Ríos, Marga Nadal, Montserrat Plana, Julià Blanco, Teresa Prat, Elia Torroella, Rafel Ramos, the HIPRA-HH-1 study group

## Abstract

**Background:** We report safety, tolerability, and immunogenicity of a recombinant protein RBD-fusion heterodimeric vaccine against SARS-CoV-2 (PHH-1V).

**Methods:** A dose-escalation, phase 1-2a, randomized clinical trial was performed in Catalonia, Spain. Each cohort had one safety sentinel that received PHH-1V vaccine of the corresponding dose, and remaining participants were randomly assigned to receive PHH-1V formulations [10µg (n=5), 20µg (n=10), 40µg (n=10)] or control BNT162b2 (n=5). Two intramuscular doses (0-21 days) were administered. Primary endpoint was solicited events 7 days after each vaccination and secondary-exploratory endpoints were humoral and cellular immunogenicity.

**Findings:** 30 young healthy adults were enrolled, thirteen were female. Vaccines were safe, well tolerated. The most common solicited events for all groups were tenderness and pain at the site of injection. The proportion of subjects with at least one reported local and/or systemic solicited adverse events (AE) after first or second vaccine dose were lowest in PHH-1V (n=21, 84%) than control group (n=5, 100%). AE were mild to moderate, and no severe AE nor AE of special interest were reported. All participants had a >4-fold change at day 35 in total binding antibodies from baseline. Variants of concern (VOC) alpha, beta, delta and gamma were evaluated using a SARS-CoV-2 pseudovirus-based neutralization assay. All groups had a significant geometric mean fold rise (p<.0001) at day 35 against all studied VOC. Similar results were obtained when a full replicative virus neutralization assay was carried out.

**Interpretation:** PHH-1V was safe, well tolerated, and induced robust humoral responses. These data support further exploration of PHH-1V in larger studies.

**Funding:** HIPRA

**ClinicalTrials.gov Identifier:** NCT05007509

**Research in context:** *Evidence before this study:* We searched PubMed up until August 1, 2021, with the terms “SARS-CoV-2”, “COVID-19” and “vaccine”. We initially identified 12,952 results but when added the terms “clinical trial” and “variants” this number decreased to 50. Of these references twelve were clinical trials, and although several vaccines were under development, and the ones that were already approved for administration in the general population described the neutralization effect to the different circulating variants of concern, we could not find any reference to a vaccine developed using variants of concern instead of ancestral Wuhan strain.

*Added value of this study:* To the best of our knowledge, our study is the first clinical trial to assess the effect as a primary series of a recombinant protein receptor-binding domain fusion heterodimer PHH-1V vaccine against SARS-CoV-2 not including the ancestral strain in its composition. This vaccine contains RBD from B·1·351 (beta) and B·1·1·7 (alpha) variants and is co-formulated with an oil-in-water adjuvant emulsion. In this first-in-human randomized clinical trial, two doses of the SARS-CoV-2 PHH-1V vaccine in a range of 10 to 40 µg/dose were safe and well-tolerated and induced robust humoral immune responses to different circulating variants of concern, including alpha (B1·1·7), beta (B·1·351), delta (B·1·617·2) and gamma (P·1). Additionally, the PHH-1V 40µg dose vaccine elicited moderated cellular immune responses, particularly to variants of concern alpha and delta.

*Implications of all the available evidence:* These findings indicate that the recombinant protein receptor-binding domain fusion heterodimer vaccine PHH-1V is safe and immunogenic. Phase 2b and Phase 3 clinical trials are ongoing to further investigate its safety and protective efficacy as heterologous booster.

## Introduction

The coronavirus disease 2019 (COVID-19) caused by the severe acute respiratory syndrome coronavirus 2 (SARS-CoV-2) is overwhelming global health and has led us to social and economic exhaustion. As a result, there has been an unprecedented effort to develop different types of vaccines against this new virus that have been remarkable^1^ and relevant in preventing severe disease and minimizing death. The great majority of these vaccines have based their antigen on a stabilized trimeric structure of the Spike glycoprotein (S) from the ancestral strain isolated in earliest identified infections. However, viral sequence evolution leading to emergence of variants has impacted on vaccine effectiveness. Moreover, there is an unmet need to cover vaccination worldwide, which is low and highly inequitable,^2,3^ while facing an increasingly transmissible virus. We need a new generation of vaccines to overcome all these challenges.

In response to this pandemic and to contribute to the solution we have launched a vaccine development program against SARS-CoV-2. PHH-1V vaccine is a SARS-CoV-2 recombinant spike (S) protein receptor binding domain (RBD) fusion heterodimer containing the B·1·351 (beta) and B·1·1·7 (alpha) variants and co-formulated with an oil-in-water emulsion adjuvant produced by the Sponsor, named SQBA. The RBD is a key functional component within the S protein that is responsible for binding SARS-CoV-2 to its cell receptor,^4^ and it is one of the main targets for neutralizing antibodies^5^. Using just the RBD region as an antigen has the advantage of focusing immunity to key protective determinants,^5,6^ which is supported as the main biomarker of protection.^7,8^ Another advantage is that these antigens can be scaled and produced faster and easier compared to the entire S protein or its subunits (S1, S2).^9^ Monomeric RBD has a limited immunogenicity possibly related to its small molecular size and the mixed forms of multiple complexes. To solve this, we have produced a highly purified RBD heterodimer with no heterologous peptide sequence added, so it is directly fused without a linker and formulated with an adjuvant to enhance the magnitude, breadth, and durability of the immune response. Other authors have disclosed homodimers with interesting results,^10,11^ but PHH-1V is the first vaccine based on heterodimers of SARS-CoV-2 variants.

Pre-clinical studies performed in mice, pigs, and non-human primates have shown that this vaccine candidate is safe and immunogenic, inducing a high titer of neutralizing antibodies against all the studied VOC of SARS-CoV-2, and promoting the activation of CD4+ and CD8+ T lymphocytes with a balanced Th1/Th2 response. Moreover, the PHH-1V vaccine has demonstrated to be efficacious against an experimental infection with SARS-CoV-2 in K18-hACE2 mice and cynomolgus monkeys. In Catalonia, Spain, we conducted a first-in-human phase 1-2a clinical trial in healthy, SARS-CoV-2 seronegative adults, younger than 40 years old, to evaluate the safety, reactogenicity and immunogenicity of 10µg, 20µg and 40µg doses of PHH-1V. Here we report the main preliminary findings.

## Methods

### Study Design and Participants

This is an ongoing first-in-human phase 1-2a dose-escalation, randomized, double-blinded, active-comparator controlled clinical trial conducted at two centers in Catalonia, Spain (Hospital Clínic de Barcelona in Barcelona city and Hospital Universitari Dr Josep Trueta in Girona city) to evaluate safety and immunogenicity of a recombinant protein RBD fusion heterodimer vaccine against SARS-CoV-2. Trial screenings started on August 16^th^ 2021, and the study is still ongoing in both centers until completing 48 weeks after the last vaccine dose. The HIPRA-HH-1 study was approved by the Spanish Agency of Medicines and Medical Devices (AEMPS) and the Research Ethics Committee (REC) of the Hospital Clínic de Barcelona and was overseen by an independent data safety monitoring board (DSMB). We present the preliminary data up to day 35 after the first vaccine dose. Eligible participants were healthy women and men adults 18 to 39 years of age who received two injections of study vaccine 21 days apart distributed in three dose-escalation cohorts 10µg, 20µg and 40µg or approved BNT162b2 vaccine as control.^12^ The age range limitation was recommended by the REC given that, when the study was evaluated other approved COVID-19 vaccines were easily available in Catalonia for anyone ≥ 40 years old.

Participants were recruited through advertising on the site’s website and social media. Subject recruitment material was reviewed and approved by the REC. At screening, all volunteers were tested for IgG antibodies against SARS-CoV-2 as evidence of previous infection and a polymerase chain reaction (PCR) was used to assess acute infections. Participants with any positive test were excluded along with participants recently exposed to persons with confirmed SARS-CoV-2 infection. Female participants of childbearing potential and men had to agree to use highly effective methods of contraception. The participant’s medical history was assessed by the study investigators in addition to reviewing clinical and laboratory findings from tests at screening following the study protocol (appendix 1). All participants provided written informed consent before enrollment in the trial.

### Randomization and masking

Participants were allocated to the dose escalation cohort according to the order that they had been preselected considering the laboratory results and subject availability. Each cohort had a safety sentinel individual that received the study vaccine of the corresponding dose. With the exception of sentinels, all participants at each dose cohort were randomly allocated to study vaccine or control vaccine in a 5:1 allocation scheme. A centralized computer-generated randomization was used, and a study independent statistician generated these randomization codes by means of the PROC PLAN of the SAS system. Randomization was centralized through the electronic Case Report Form (eCRF) created using the Elsevier Macro® system. This system is regulatory compliant (ICH GCP and FDA 21 CRF Part11). More participants were allocated to the higher dose groups based on preclinical experience. Study investigators and participants were both blinded, only study staff responsible for preparing and administering the vaccine were unblinded and were not involved in assessment of study data. Syringes were masked using opaque labels since study and control vaccines were visually different.

### Procedures

Initially, the study vaccine was based on sequence of the SARS-CoV-2 strain first detected in Wuhan, but due to the rapid spread of new variants of concern (VOC) around the world, the sponsor decided to develop a new antigen candidate, based on the same Chinese Hamster Ovary (CHO) cells platform technology, considering variants B·1·351 and B·1·1·7. This new candidate also elicited a cross-reactive response and neutralization against heterologous pseudoviruses Wuhan strain, P·1 and B·1·617·2 in pre-clinical studies.^13^ Different adjuvants, alone or combined, were assayed. Based on non-clinical studies, the oil-in-water emulsion adjuvant SQBA was selected. Study vaccines were packed as single vials with 0.5ml emulsion ready to use and were stored at 2 - 8ºC. Due to AEMPS and REC recommendations we included a comparator control group, only considering safety assessment. This comparator group was an approved mRNA vaccine, and its selection was made considering the similar posology. The study vaccine was developed by researchers at HIPRA (Amer, Girona, Spain) and they were also involved in discussions of the trial design, provided the vaccine candidate and, as part of the writing group, contributed to drafting the manuscript.

All vaccines were administered as a single intramuscular injection into the deltoid muscle at days 0 and 21. Each sentinel individual in each dose cohort was monitored by phone for 24 and 48 hours after the first administration. Early safety data from sentinels was reviewed by an Internal Review Committee (IRC) before including the remaining participants of each group. Further participants in the same cohort were randomized to receive either study vaccine or control vaccine and were distributed in small groups of five-to-six participants per day and safety data was monitored for 24hrs. After 48-72hrs of the last vaccine administered in each dose cohort, DSMB assessed if any clinically significant adverse events occurred and if no halting rules were met, study vaccine dose was escalated. All participants were observed for 60-120 minutes after each vaccine dose on site. Participants from the same cohort received the vaccine with an interval of 60 minutes between them. Details of the trial design, conduct and analyses are provided in the protocol (appendix 1).

During the first seven days after each vaccination, any solicited local and systemic adverse events (AE) were self-reported by participants daily on the diary cards and verified by the investigator during the scheduled visit. Unsolicited local and systemic AE occurring within the 28 days after each vaccination were reported by participants during scheduled follow-up visits or by any preferred method if occurring before. Any unsolicited AE, serious AE (SAE), AE of special interest (AESI) or medically attended AE (MAAE) occurring within 48 weeks after receiving the second vaccination dose was monitored, and follow-up is still ongoing. Solicited local AE included pain, tenderness, erythema, and swelling; and solicited systemic AE included fever, nausea/vomiting, diarrhoea, headache, fatigue and myalgia. Laboratory safety tests including routine blood and serum chemistry were done to assess any short- and long-term toxicity after vaccination. AE and changes in laboratory tests were assessed according to the Guidance for Industry, Toxicity Grading Scale for Healthy Adult and Adolescent Volunteers Enrolled in Preventive Vaccine Clinical Trials by the U.S. Department of Health and Human Services, FDA, Center for Biologics Evaluation and Research (September 2007).

Blood samples for safety assessment were collected at screening, seven and 28 days after each vaccine dose and at weeks 12, 24 and 48 after the second vaccine dose. To assess the immunogenicity, blood samples were collected at screening, 21 and 35 days after the first dose and at weeks 12, 24 and 48 after the second vaccination. Serum samples were collected to evaluate binding and neutralizing antibodies. Peripheral blood mononuclear cells (PBMCs) were collected to assess specific T-cell responses. The in-vitro quantitative determination of binding antibodies (including IgG) against RBD was assessed by ELISA (Elecsys SARS-CoV-2, Roche Diagnostics GmbH, D-68305 Manheim). The assay used a recombinant protein representing the RBD in a double-antigen sandwich assay format, which favors detection of high affinity antibodies against this virus and expressed as the geometric mean titers (GMTs). Pseudovirus-based Neutralization Assay (PBNA) was assessed for the alpha, beta, delta and gamma VOC. The validated PBNA consists of the use of HIV-based pseudoviruses that express the corresponding S protein of SARS-CoV-2 VOC along with luciferase as a reporter gene. A validated virus neutralization assay (VNA) was performed using an Alpha SARS-CoV-2 isolate sequenced and deposited in GISAID (ID: EPI_ISL_1663569). Viral-induced cytopathic effect of this VOC preincubated with serial dilutions of serum from vaccinated individuals was measured on Vero E6 cells using the CellTiter Glo Luciferase Cell Viability Assay (Promega) (appendix 2). T-cell mediated immunity was assessed by IFN-γ-based ELISPOT and intracellular cytokine staining (ICS).

In December 2021, due to the restrictions imposed to the Catalan population during the 6^th^ covid-19 wave and, in an intent to minimize the interference in daily-life activities of the study participants, the study was single-unblinded and a non-travel Catalan vaccination certificate was issued for participants allocated to study vaccine and a standard Spanish vaccination certificate for the control group. Participants signed a confidentiality agreement not to disclose the allocation group to investigators who were to continue the vaccine safety evaluation. Participants were informed about safety and immunogenicity results obtained so far in the study and a safety-oriented recommendation was given as for receiving any available approved covid-19 vaccine. All individuals were asked to continue their participation in the study until completion. All these changes were approved by the AEMPS and the REC.

### Outcomes

The primary endpoint was safety and tolerability of study vaccine, and it was assessed as solicited local and systemic reactogenicity adverse events within 7 days following each vaccination and unsolicited local and systemic reactogenicity adverse events within 28 days following each vaccination. Serious adverse events (SAE), adverse events of special interest (AESI) and medically attended adverse events (MAAE) related to study vaccines will be monitored throughout the study duration. Secondary endpoints related to immunogenicity were defined as antibody neutralization measured as IC50 or ID50 and expressed as geometric mean titers (GMT) and geometric mean fold rise (GMFR) from baseline to days 21, 35 and weeks 12, 24, 48; binding antibodies titer measured as GMT and GMRF from baseline to days 21, 35 and weeks 12, 24, 48 and T-cell mediated response measured by ELISpot and ICS at baseline and day 35. Other exploratory endpoints related to COVID-19 cases were assessed. Seroconversion were defined in two ways: as ≥ 4-fold change in binding antibody titer and a titer above 0.8 U/mL from baseline to days 21 and 35.

### Statistical Analysis

Main population analysis, since this Phase 1-2a clinical trial is not confirmatory, it is not possible to justify the sample size numerically in the usual terms of confirmatory trials.

However, the sample size seems reasonable in this exploratory context. With a sample size of n=10 for each 20µg or 40µg dose cohort, the probability of observing at least one AE with a prevalence rate of ≥ 10% is 65.1%. In the 10µg dose cohort, with a sample size of 10, this probability is 41.0%. For participants in all active groups, n=25, this probability will be 92.8%. These calculations were performed with the nQuery Advisor program version 7.0.Endpoints related to the primary safety outcomes, number of solicited and unsolicited AEs described previously, as well binary variables related to the immunogenicity, proportion of seroconverted subjects, were described by a proportion and 95% confidence interval (95% CI) using exact binomial-based methods, the Clopper-Pearson method, and the Clopper-Pearson method.^14^ Quantitative results related to immunogenicity and T-cell measurements were analyzed, on previously log-transformed data, using restricted maximum likelihood (REML)-based repeated measures approach (MMRM: Mixed Models for Repeated Measurements). Analyses will include the fixed, categorical effects of group, visit, and group-by-visit interaction. A common unstructured structure were used to model the within-patient correlation. The Kenward-Roger approximation will be used to estimate denominator degrees of freedom.^15^ Estimation of effects between and within group were assessed by the ratio and this 95% CI between geometric means. Since this is an exploratory Phase I trial with no formal interim analysis for early study termination, no alpha adjustments are needed to maintain the type-I error.^16^ The statistical software used to analyze all data was SAS version 9.4 (SAS Institute Inc., Cary, NC).

Role of the funding source: The funders of the study were involved in study design, data collection, data analysis, data interpretation, writing of the report, and the decision to submit the manuscript for publication.

## Results

### Trial population

Between August 16 and September two, 2021, 51 healthy adults were screened and after the eligibility assessment 21 were excluded. Of the 30 participants included, three were allocated as sentinels at each PHH-1V dose group and the remaining 27 were randomly assigned into four groups to receive PHH-1V 10µg (n=4), PHH-1V 20µg (n=9), PHH-1V 40µg (n=9) or control vaccine BNT162b2 (n=5). Thirteen participants were female (43·3%), the mean age was 27·7 (SD 4·91) and most of them were Hispanic (n=29, 96·7%). All participants completed the two-dose scheme and attended the day 35 visit as scheduled. Subject disposition is shown in figure 1. The trial is ongoing at the time of writing this manuscript and it is expected to be ended in September 2022 when completing 48 weeks follow-up after the last vaccine dose for all participants.

**Figure 1.**
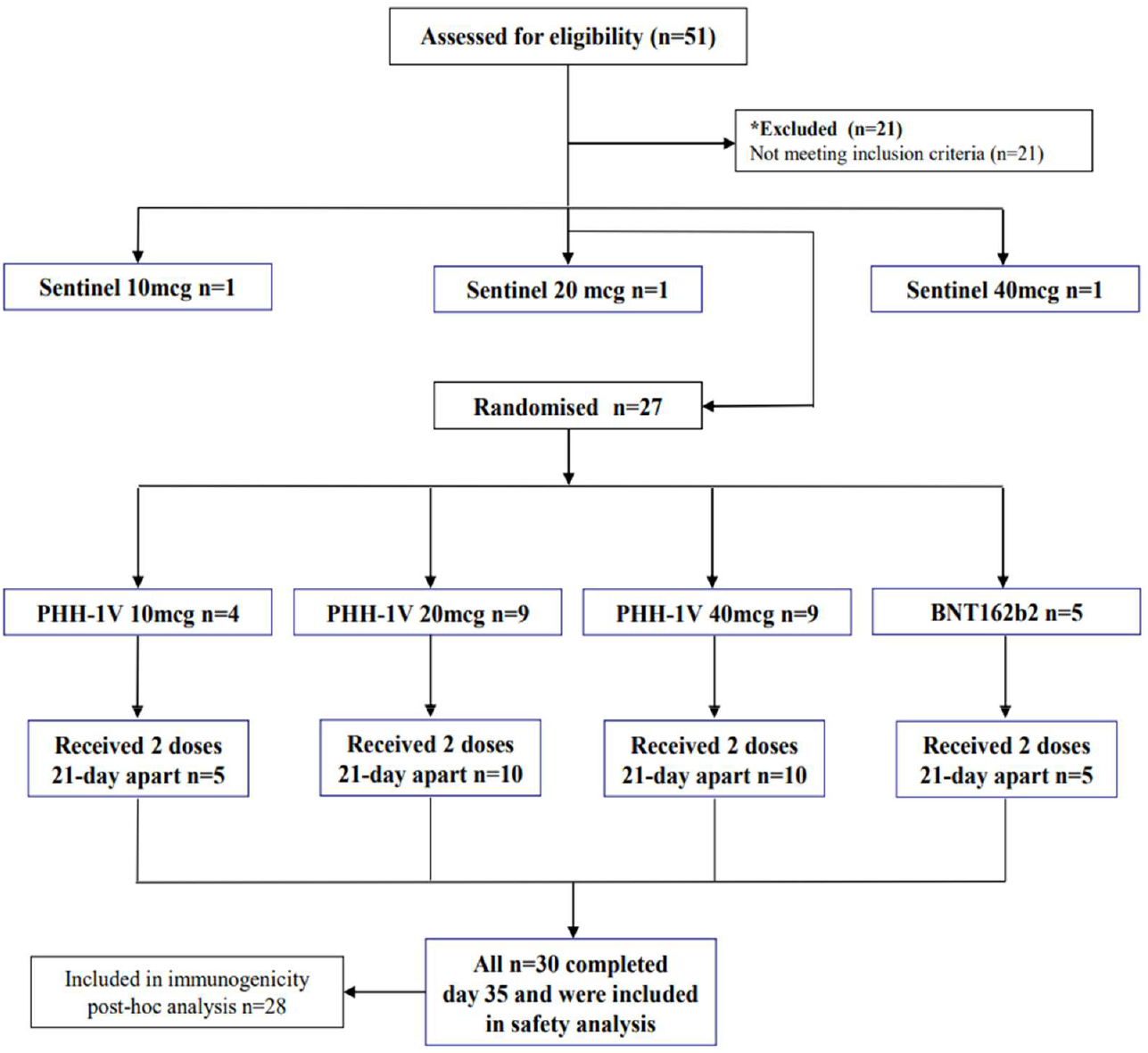
Subject disposition according to Consort guidelines.

### Safety outcomes

Vaccines were safe and well tolerated. All solicited adverse events reported were mild to moderate (grade 1 and 2), transient and resolved within the reporting period. Twenty-six subjects (86·7%; 95%IC: 69·3% - 96·2%) referred to having at least one solicited AE within seven days after first or second vaccine dose. Twenty-four individuals (80·0%; 95%IC: 61·4% - 92·3%) after the first vaccination and 19 (63·3%; (95%CI: 43·9% - 80·1%) after the second one. The most common solicited events for all groups were tenderness and pain at the site of injection followed by headache and fatigue (Figure 2). Two participants from the BNT162b2 control group had fever, defined as temperature ≥ 38ºC, within seven days after the second vaccination. No prophylactic treatment was prescribed. Eight participants, four in PHH-1V 20µg, one in PHH-1V 40µg and three in the control group, took occasional paracetamol and other painkillers within seven days after vaccination. From day 0 through day 28 following vaccination 23 (76·7%; 95%CI: 57·7% - 90·1%) participants reported 61 AE of which 14 (46·7%; 95%CI: 28·3% - 65·7%) participants reported 35 were considered related to the vaccines (PHH-1V 10µg n=8, 20µg n=13, 40µg n=12, BNT162b2 n=2). The most frequent unsolicited events described were related to the respiratory tract (n=8); in all these cases a SARS-CoV-2 PCR was performed, and COVID-19 infection was discarded. There were four reported MAAE (for 3 subjects) of which three (in 2 subjects) corresponded to the PHH-1V 20µg group and one to the PHH-1V 40µg group; only two were considered as possibly related to the vaccine. There were no SAE nor AESI reported until day 35 follow-up. No cases of SARS-CoV-2 infections or COVID-19 were reported until day 35 follow-up.

**Figure 2.**
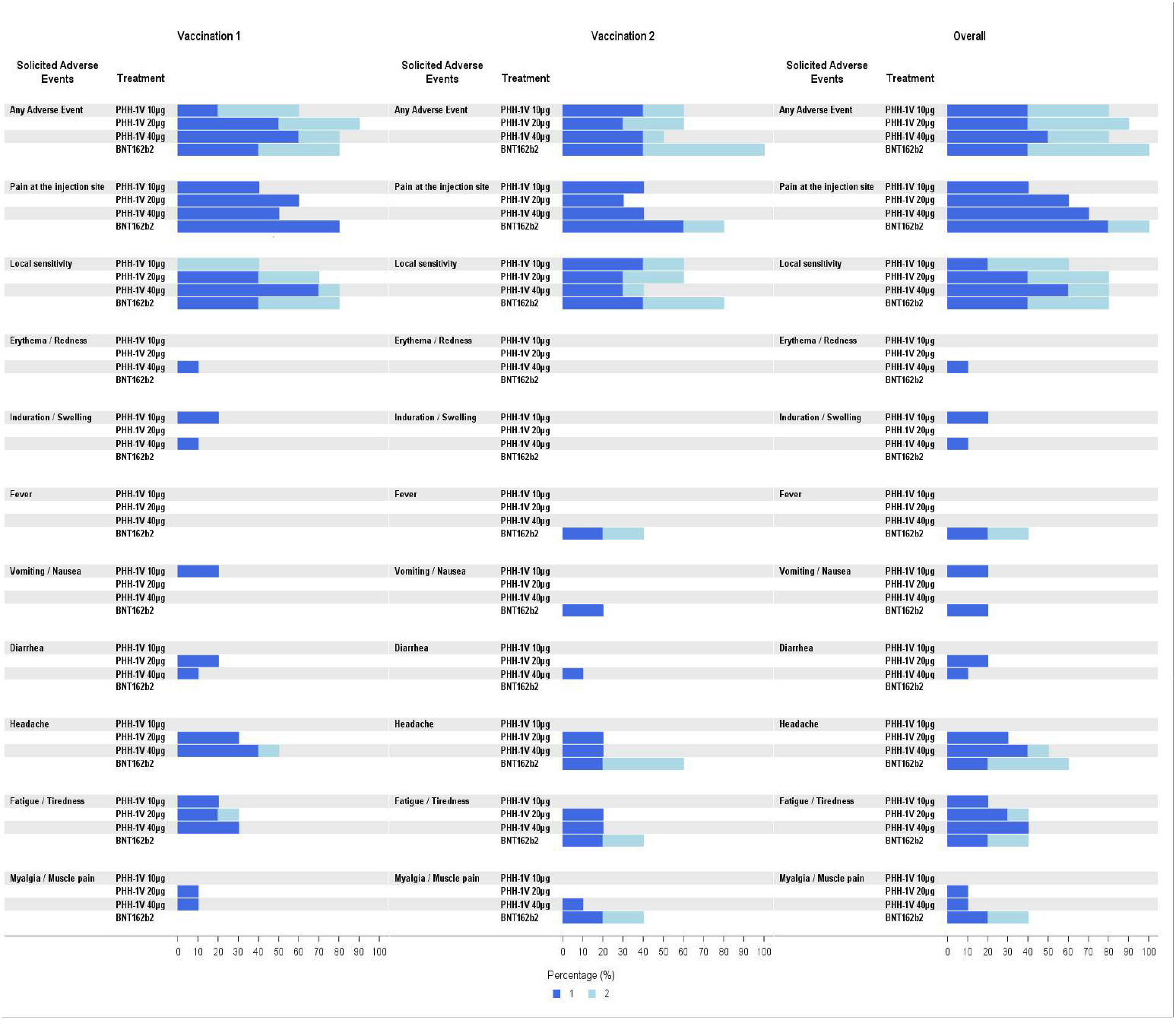
Solicited Adverse Events (AEs) reported by percentage within 7 days after each vaccination and overall, by group. These AE were reported by participants in their diaries as well as during follow-up visits.

### Immunogenicity outcomes

Total binding antibodies titers were assessed by ELISA. At baseline, all participants but two had binding antibodies titers lower than the limit of detection (0·8 U/ml), one female in the PHH-1V 40µg group (8·4 U/ml) and another female in the control group (8·77 U/ml). By day 21, twenty-nine (97%) participants had seroconverted, and binding antibodies had >4-fold change from baseline. The only participant that had undetectable levels had received a dose of PHH-1V 10µg. At day 35 all participants had seroconverted and 100% had >4-fold change (Figure 3). Binding antibodies GMT were 11·31 (95% CI 1·40 – 91·66) at day 21 and 334·18 (95% CI 118·71 - 940·76) at day 35 for PHH-1V 10µg [GMFR 1670·88 (95% IC 708·99 - 3937·80) p<0.0001]; 6·93 (95% CI 1·58 – 30·42) at day 21 and 358·15 (95% CI 172·27 - 744·57) at day 35 for PHH-1V 20µg [GMFR 1790·73 (95% CI 976·72 - 3283·17) p <0·0001]; 26·09 (95% CI 5·94 - 114·56) at day 21 and 827·61 (95% CI 398·09 - 1720·57) at day 35 for PHH-1V 40µg [GMFR 2847·56 (95% CI 1553·14 - 5220·77) p<0·0001]; and 202·25 (95% CI 24·96 - 1638·91) at day 21 and 2821·43 (95% CI 1002·23 - 7942·75) at day 35 for control vaccine [GMFR 6622·85 (95% CI 2810·2 - 15608·22) p<0·0001].

**Figure 3.**
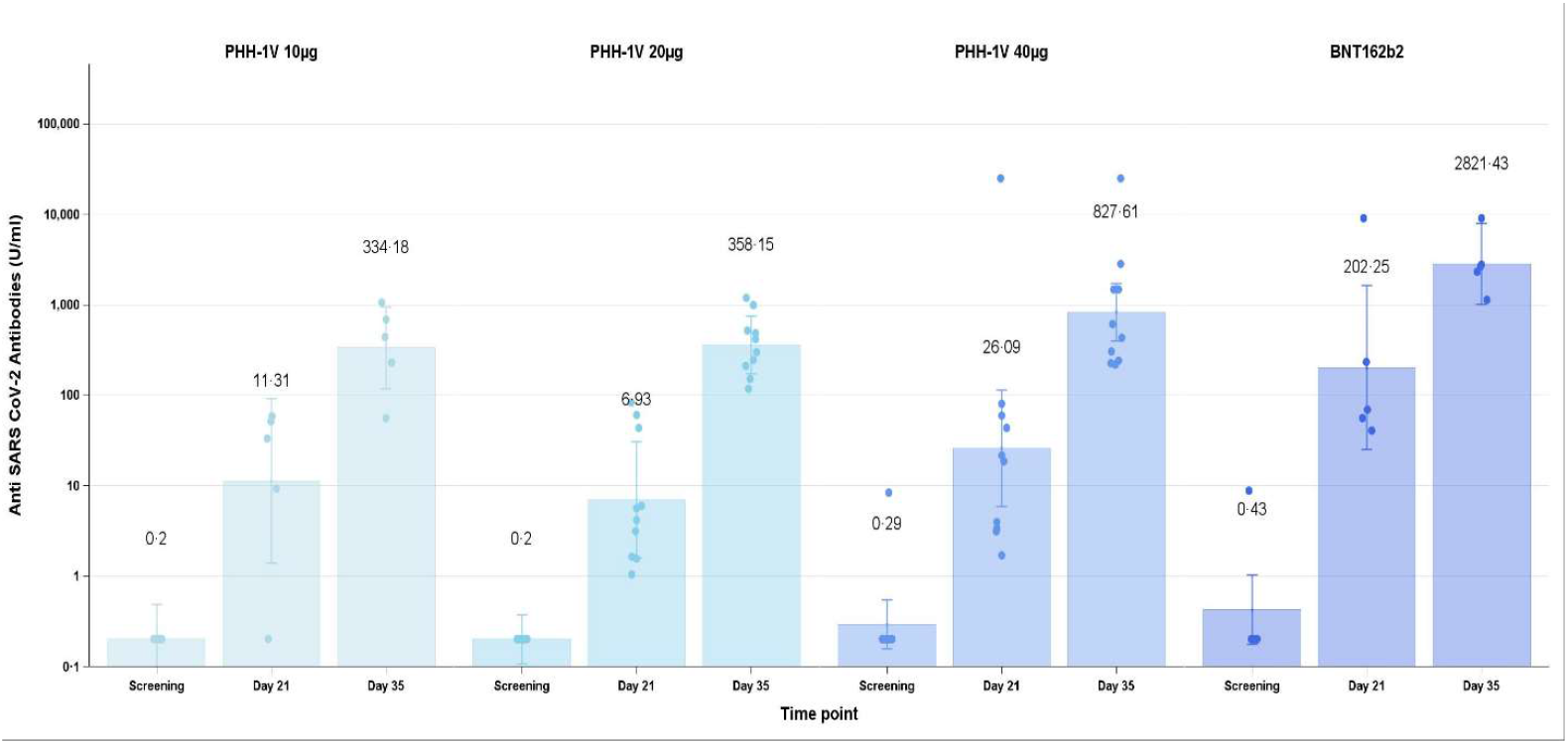
Total binding antibodies titers for all groups. Measured total binding antibodies by ELISA at screening, 21 days after the first vaccination (day 21) and 14 days after the second vaccination (day 35). Binding is expressed as geometric mean titers (GMT)

Neutralization assays were conducted using wild-type SARS-CoV-2 and SARS-CoV-2 pseudoviruses. The correlation between these two techniques was excellent for the alpha variant (r=0·93, p<0·0001). The pseudovirus-based neutralization assay (PBNA) analyzed VOC present at that moment (alpha, beta, gamma and delta). The participant in the PHH-1V 40µg group with detectable binding antibodies at baseline also had detectable neutralizing antibodies against all VOC by PBNA at the same time-point; this was interpreted as a cross-reaction to other coronaviruses or a previous SARS-CoV-2 asymptomatic infection with a false-negative IgG antibody detection at the screening.

On day 21, twenty-four (80%) participants had detectable titers of neutralizing antibodies by PBNA to all the studied VOC as follows: PHH-1V 10µg n=4, PHH-1V 20µg n=10, PHH-1V 40µg n=10, BNT162b2 n=5. All participants had detectable neutralizing antibodies for all VOC at day 35.

Neutralization GMT and GMFR for all VOC can be seen in table 1 and figure 4. Considering that the two participants that had baseline antibody titers above the limit of detection could have had a previous SARS-CoV-2 infection, we also conducted a post-hoc analysis excluding them. This additional analysis did not modify the main conclusions of the study (data not shown).

**Table 1.**
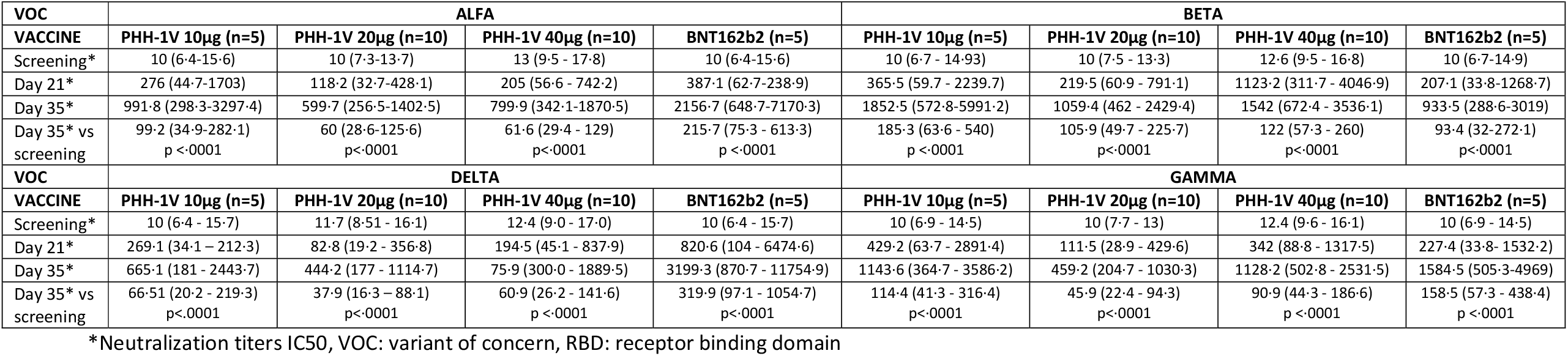
Neutralization titres measured by pseudovirus-based neutralization assay (PBNA) by time-point, vaccine group and variant of concern assessed.

**Figure 4.**
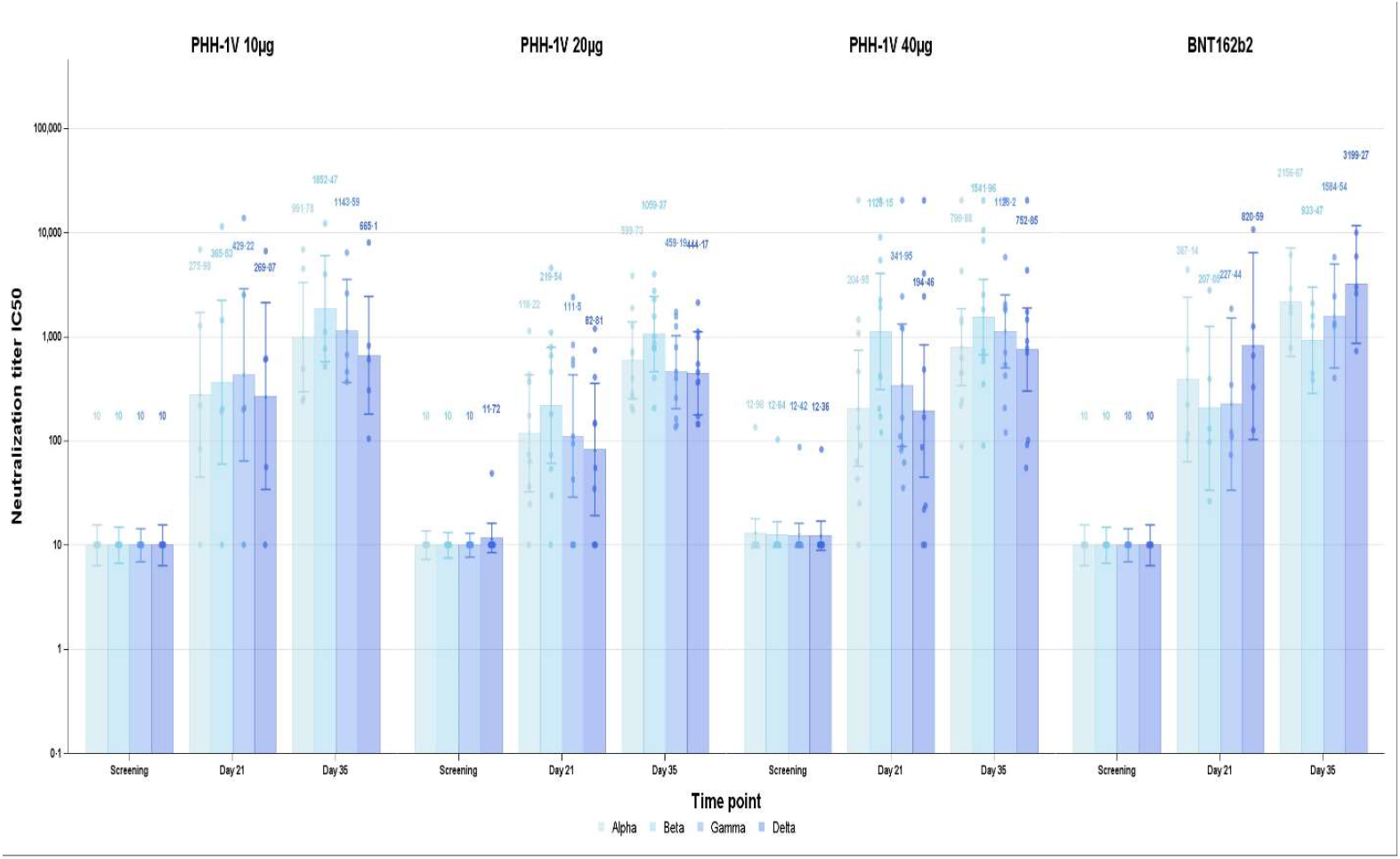
Neutralizing antibodies titers for all groups. Measured responses against alpha (B·1·1·7), beta (B·1·351), delta (B·1·617·2) and, gamma (P·1) variants of concern by pseudo-virus neutralization assay (PNBA) at screening, 21 days after the first vaccination (day 21) and 14 days after the second vaccination (day 35). Neutralization is represented as IC50.

T-cell mediated immunogenicity was assessed using IFN-γ ELISPOT. For this analysis we used six peptide pools of overlapping SAR-CoV-2 peptides each encompassing the SARS-CoV-2 regions S, RBD Wuhan, RBD 1·1·7 (alpha), RBD B·1·351 (beta) and RBD B·1·617·1 (delta) (JPT Peptide Technologies GmbH, Germany). At day 35 results showed that vaccination with PHH-1V 40µg induced a specific T cell response, with a significant IFN-ɣ production after re-stimulation in vitro with RBD peptides from alpha [2.07 SFC/106 PBMC (95% CI 1·04-4·14) p= 0·0398] and delta [2·99 SFC (95% CI 1·2-7·42) p= 0·0202] SARS-CoV-2 variants compared with baseline. Changes in the T-cell responses by groups and between different timings and VOC are exposed in table 2 and appendix 3. We had technical problems when analyzing samples by ICS, thus we exclude it from our report.

**Table 2.** Total SARS-CoV-2 specific T-cell responses measured by IFN-γ ELISPOT by time-point, vaccine group and variant of concern assessed.

| VOC | ALFA |  |  |  | BETA |  |  |  |
| --- | --- | --- | --- | --- | --- | --- | --- | --- |
| VACCINE | PHH-1V 10µg (n=5) | PHH-1V 20µg (n=10) | PHH-1V 40µg (n=10) | BNT162b2 (n=5) | PHH-1V 10µg (n=5) | PHH-1V 20µg (n=10) | PHH-1V 40µg (n=10) | BNT162b2 (n=5) |
| Screening* | 4.92 (2.31 - 10.48) | 2.38 (1.39 - 4.06) | 1.25 (0.73 - 2.13) | 2.42 (1.06 - 5.52) | 4.36 (2.16 - 8.83) | 2.19 (1.33 - 3.61) | 1.88 (1.14 - 3.09) | 1.16 (0.54 - 2.49) |
| Day 21* | 1.43 (0.57 - 3.61) | 1.67 (0.87 - 3.21) | 3.1 (1.61 - 5.96) | 5.69 (2.25 - 14.36) | 2.27 (1.09 - 4.7) | 1.17 (0.7 - 1.97) | 4.06 (2.43 - 6.81) | 7.21 (3.48 - 14.94) |
| Day 35* | 3.19 (1.26 - 8.05) | 2.15 (1.12 - 4.15) | 2.58 (1.34 - 4.97) | 18.62 (7.38 – 47) | 2.86 (1.1 - 7.45) | 2.97 (1.51 - 5.85) | 2.33 (1.19 - 4.59) | 19.4 (7.45 - 50.5) |
| Day 35 vs screening | 0.65 (0.24 - 1.73)<br>p=0.3715 | 0.91 (0.45 - 1.81)<br>p=0.7733 | 2.07 (1.04 - 4.14)<br>p=0.0398 | 7.69 (2.74 - 21.6)<br>p=0.0004 | 0.66 (0.2 - 2.2)<br>p=0.4796 | 1.35 (0.58 - 3.19)<br>p=0.4721 | 1.24 (0.53 - 2.93)<br>p=0.6040 | 16.77 (4.81 - 58.43)<br>p<0.0001 |
| VOC | DELTA |  |  |  | RBD PEPTIDE MIX |  |  |  |
| VACCINE | PHH-1V 10µg (n=5) | PHH-1V 20µg (n=10) | PHH-1V 40µg (n=10) | BNT162b2 (n=5) | PHH-1V 10µg (n=5) | PHH-1V 20µg (n=10) | PHH-1V 40µg (n=10) | BNT162b2 (n=5) |
| Screening* | 5.04 (2.46 - 10.33) | 1.62 (0.98 - 2.7) | 1.73 (1.04 - 2.88) | 1.11 (0.51 - 2.4) | 7.22 (3.24 – 16.09) | 2.93 (1.66 – 5.16) | 2.35 (1.33 – 4.14) | 1.7 (0.7 – 4.13) |
| Day 21* | 2.17 (0.81 - 5.78) | 1.37 (0.69 - 2.75) | 2.89 (1.44 - 5.78) | 6.37 (2.39 - 16.98) | 2.49 (0.95 – 6.51) | 1.85 (0.94 – 3.66) | 2.73 (1.38 – 5.38) | 4.6 (1.76 – 12.03) |
| Day 35* | 5.28 (2.04 - 13.68) | 2.55 (1.3 - 4.99) | 5.17 (2.64 - 10.14) | 20.72 (8 - 53.69) | 6.79 (2.67 – 17.23) | 3.47 (1.79 – 6.7) | 3.92 (2.03 – 7.58) | 16.02 (6.31 - 40.66) |
| Day 35 vs screening | 1.05 (0.29 - 3.79)<br>p=0.9411 | 1.57 (0.63 - 3.89)<br>p=0.3183 | 2.99 (1.2 - 7.42)<br>p=0.0202 | 18.68 (5 - 69.77)<br>p=0.0001 | 0.94 (0.33 – 2.71)<br>p=0.9047 | 1.18 (0.56 – 2.5)<br>p=0.6464 | 1.67 (0.79 – 3.52)<br>p=0.1720 | 9.45 (3.06 – 29.15)<br>p=0.0004 |

\*mean spot forming cells / 106 PBMC (95% CI), VOC: variant of concern, RBD: receptor binding domain

## Discussion

Vaccination remains an essential component of the approach to fighting against the ongoing pandemic, but limited supply, storage requirements and vaccine hesitancy have restricted their global impact. As we face the rapid emergence and spread of new variants, existing vaccines are losing their efficacy, so developing a new generation of multivalent vaccines, which provide cross-strain protection seems a reasonable strategy to pursue.

Adjuvanted protein-based vaccines, using a more traditional inexpensive technology and already in widespread use for other diseases, have a lot to offer and could change the pandemic.^17^ PHH-1V vaccine has shown to be safe and well-tolerated in healthy young adults. Furthermore, the two-dose regimen induced a robust binding and neutralizing antibodies against the different VOC at all PHH-1V vaccine concentrations and elicited a moderate specific T-cell response at the highest dose of 40ug.

PHH-1V vaccine safety profile is comparable to other vaccines using the same platform technologies.^12,18^ There were no severe adverse events or study withdrawals. The most frequent AE were local pain and tenderness and most of them reported after the first dose of PHH-1V vaccine contrarily to the control group in which most of AE were reported after the second dose, as has already been described.^19^ Fever was only described in the control group and although no prophylactic antipyretic was prescribed, proportionally more participants in the control group had to take occasional paracetamol or non-steroidal painkillers after vaccinations. Overall, PHH-1V has a good safety profile, we need to continue this evaluation at a larger scale to corroborate this. Interesting to point out that participants as well as investigators were blinded during these assessments, which had somehow limited the bias. The study was conducted at a time of decreasing daily diagnoses of COVID-19, and there were no cases reported until the study time point described in this manuscript. All study participants had seroconverted after 14 days from the second dose and at that same time point, they all had a >4-fold increase in neutralizing antibodies titers. It is presumed that neutralizing antibodies are associated with protection.^20,21^ PHH-1V 10µg was fairly immunogenic but at higher doses there was a robust increase in neutralizing activity, yet evident after the first dose and significantly enhanced after the second, similar as to what has been showed by other SARS-CoV-2 protein vaccines.^22,23^ In the different clinical trials evaluating the efficacy of SARS-CoV-2 vaccines that nowadays are widely used, neutralization GMT were assessed and are also considered as surrogate markers of protection. Unfortunately, different analytical methods were used, and the results vary for the different vaccines, therefore indirect comparison between vaccines may not be reliable.^24,25^ In any case, this PHH-1V neutralization response could be associated with a protective effect against SARS-CoV-2 infection.

This study has limitations. The interpretation of the results is limited due the small sample size and the short follow-up until the moment this manuscript was written. Because this is an interim report, we are not reporting any data on long-term safety outcomes and the results obtained do not permit efficacy assessments. All the participants in this phase 1-2a study were very young, therefore, safety, tolerability and immunogenicity are under evaluation in a larger phase 2b and 3 clinical trials with a representative older population. Based on the safety and immunogenicity data obtained through day 35, the two-dose regimen of 40µg PHH-1V was determined to be the optimal dose scheme to continue with the development plan of the PHH-1V vaccine.

Due to the evolution of the ongoing pandemic and successful vaccine coverage in Europe with first generation vaccines, PHH-1V will be further evaluated as a booster.

In conclusion, we found that this recombinant protein RBD fusion heterodimer vaccine PHH-1V is safe, well tolerated, and immunogenic in healthy, young people. PHH-1V is a promising vaccine candidate that would make a valuable addition to the global COVID-19 response. We will continue further investigation to demonstrate its relevance and understand the best possible way to contribute to world-wide health with this vaccine.

## Data Availability

Data collected for the study is available under request to the Sponsor.

## Authors contributions

LL, JP, JAA, TP, GD, JR, and ET designed the study. LF, AP, CG, AB, MP, AS, MN and RR contributed to the study design. JP, KE, JC, and CG contributed to data management. LL, JN, RMLL, MRA, ACC and RR recruited and follow up participants. LFIE, EP, DRR, AP, BT, JMB, NIU, MP, CP and JB performed the experiments. JR and GS undertook the statistical analysis. LL, JP, LF, JB, NIU, JR, GD and MP drafted the manuscript. All authors participated in study analyses and revised the manuscript critically for important scientific and intellectual content. All authors approved the final version and made the decision to submit for publication.

## Data sharing

Data collected for the study is available under request to the Sponsor.

## Acknowledgements

We would like to thank all the hundreds of persons that volunteered to collaborate in this study, and very especially to the participants.

We are indebted to the HCB-IDIBAPS and IDIBGI Biobanks, both integrated in the Spanish National Biobanks Network, for the biological human samples and data procurement.

From HIPRA to Glòria Pujol and Eduard Fossas for their assistance in the revision of the manuscript; Fiorella Gallo, Núria Fuentes and Miriam Oria for the ELISA analysis; Clara Panosa, Thais Pentinat and Ester Puigvert for their assistance in the production of the vaccine; Jordi Palmada and Eva Pol for carrying out manufacturing controls, and last but not least, to all HIPRA employees who in one way or another have contributed to making this project a reality.

## References

1. World Health Organization. COVID-19 vaccine tracker and landscape. Accessed December 16, 2021. https://www.who.int/publications/m/item/draft-landscape-of-covid-19-candidate-vaccines

2. Looi MK. The world according to covid vaccine coverage. BMJ. Published online November 11, 2021:2732. doi:10.1136/bmj.n2732

3. Global Change Data Lab. Our World In Data. Coronavirus (COVID-19) Vaccinations. https://ourworldindata.org/covid-vaccinations.

4. Lan J, Ge J, Yu J, et al. Structure of the SARS-CoV-2 spike receptor-binding domain bound to the ACE2 receptor. Nature. 2020;581(7807):215–220. doi:10.1038/s41586-020-2180-5

5. Greaney AJ, Starr TN, Barnes CO, et al. Mapping mutations to the SARS-CoV-2 RBD that escape binding by different classes of antibodies. Nature Communications. 2021;12(1):4196. doi:10.1038/s41467-021-24435-8

6. Greaney AJ, Loes AN, Gentles LE, et al. Antibodies elicited by mRNA-1273 vaccination bind more broadly to the receptor binding domain than do those from SARS-CoV-2 infection. Science Translational Medicine. 2021;13(600). doi:10.1126/scitranslmed.abi9915

7. Kleanthous H, Silverman JM, Makar KW, Yoon IK, Jackson N, Vaughn DW. Scientific rationale for developing potent RBD-based vaccines targeting COVID-19. npj Vaccines. 2021;6(1):128. doi:10.1038/s41541-021-00393-6

8. Khoury DS, Cromer D, Reynaldi A, et al. Neutralizing antibody levels are highly predictive of immune protection from symptomatic SARS-CoV-2 infection. Nature Medicine. 2021;27(7):1205–1211. doi:10.1038/s41591-021-01377-8

9. Dai L, Gao GF. Viral targets for vaccines against COVID-19. Nature Reviews Immunology. 2021;21(2):73–82. doi:10.1038/s41577-020-00480-0

10. Dai L, Zheng T, Xu K, et al. A Universal Design of Betacoronavirus Vaccines against COVID-19, MERS, and SARS. Cell. 2020;182(3):722-733.e11. doi:10.1016/j.cell.2020.06.035

11. Yang S, Li Y, Dai L, et al. Safety and immunogenicity of a recombinant tandem-repeat dimeric RBD-based protein subunit vaccine (ZF2001) against COVID-19 in adults: two randomised, double-blind, placebo-controlled, phase 1 and 2 trials. The Lancet Infectious Diseases. 2021;21(8):1107–1119. doi:10.1016/S1473-3099(21)00127-4

12. European Medicine Agency. Science Medicine Health. Cominarty. https://www.ema.europa.eu/en/medicines/human/EPAR/comirnaty.

13. Barreiro A, Prenafeta A, Bech-Sabat G, et al. Preclinical efficacy, safety, and immunogenicity of PHH-1V, a second-generation COVID-19 vaccine candidate based on a novel recombinant RBD fusion heterodimer of SARS-CoV-2. bioRxiv. Published online 2021.

14. Agresti A. Categorical Data Analysis. 2nd Edition, John Wiley & Sons, Inc; New York; 2002.

15. Mallinckrod CH, Lane PW, Schnell D, Peng Y, Mancuso JP. Recommendations for the Primary Analysis of Continuous Endpoints in Longitudinal Clinical Trials. Ther Innov Regul Sci. 2008. doi:10.1177/009286150804200402

16. CPMP/EWP/908/99. Points to Consider on Multiplicity issues in Clinical Trials. URL: http://www.ema.europa.eu/ema/index.jsp?curl=pages/regulation/general/general_content_001220.jsp&mid=WC0b01ac05807d91a4, last accessed: 14-July-2022.

17. Dolgin E. How protein-based COVID vaccines could change the pandemic. Nature. 2021;599(7885):359–360. doi:10.1038/d41586-021-03025-0

18. Keech C, Albert G, Cho I, et al. Phase 1–2 Trial of a SARS-CoV-2 Recombinant Spike Protein Nanoparticle Vaccine. New England Journal of Medicine. 2020;383(24):2320–2332. doi:10.1056/NEJMoa2026920

19. Polack FP, Thomas SJ, Kitchin N, et al. Safety and Efficacy of the BNT162b2 mRNA Covid-19 Vaccine. New England Journal of Medicine. Published online 2020. doi:10.1056/nejmoa2034577

20. Khoury DS, Cromer D, Reynaldi A, et al. Neutralizing antibody levels are highly predictive of immune protection from symptomatic SARS-CoV-2 infection. Nature Medicine. 2021;27(7):1205–1211. doi:10.1038/s41591-021-01377-8

21. Abbasi J. Homing In On a SARS-CoV-2 Correlate of Protection. JAMA. 2022;327(2):115. doi:10.1001/jama.2021.24117

22. Richmond P, Hatchuel L, Dong M, et al. Safety and immunogenicity of S-Trimer (SCB-2019), a protein subunit vaccine candidate for COVID-19 in healthy adults: a phase 1, randomised, double-blind, placebo-controlled trial. The Lancet. 2021;397(10275):682–694. doi:10.1016/S0140-6736(21)00241-5

23. Sridhar S, Joaquin A, Bonaparte MI, et al. Safety and immunogenicity of an AS03-adjuvanted SARS-CoV-2 recombinant protein vaccine (CoV2 preS dTM) in healthy adults: interim findings from a phase 2, randomised, dose-finding, multicentre study. The Lancet Infectious Diseases. Published online January 2022. doi:10.1016/S1473-3099(21)00764-7

24. He Q, Mao Q, Zhang J, et al. COVID-19 Vaccines: Current Understanding on Immunogenicity, Safety, and Further Considerations. Frontiers in Immunology. 2021;12. doi:10.3389/fimmu.2021.669339

25. Krammer F. A correlate of protection for SARS-CoV-2 vaccines is urgently needed. Nature Medicine. 2021;27(7):1147–1148. doi:10.1038/s41591-021-01432-4

