## Appendix for "Safety and Immunogenicity of a Recombinant Protein RBD Fusion Heterodimer Vaccine against SARS-CoV-2: preliminary results of a phase 1-2a dose-escalating, randomized, double-blind clinical trial"

**1-Study protocol**

Link to protocol

**2-Supplementary methods**

**Biosafety Approval.**

The biologic biosafety committee of the Research Institute Germans Trias i Pujol approved the execution of SARS-CoV-2 experiments at the BSL3 laboratory of the Center for Bioimaging and Comparative Medicine (CSB-20-015-M3).

**Cell culture and Viral isolation and titration.**

Vero E6 cells (ATCC CRL-1586) were cultured in Dulbecco’s modified Eagle medium (Invitrogen) supplemented with 10% fetal bovine serum (FBS; Invitrogen), 100 U/ml penicillin, 100 μg/ml streptomycin, (all from Invitrogen).

SARS-CoV-2 was isolated from a nasopharyngeal swab collected in January 2021 in Spain in Vero E6 cells as described in (1). The virus stock was prepared collecting the supernatant from Vero E6 and sequenced as detailed in (1). Genomic sequence was deposited at GISAID repository (http:// gisaid.org) with accession number EPI_ISL_1663567. Viral stocks were propagated in Vero E6 cells for two passages and titrated in 10-fold serial dilutions to calculate the TCID_50_ per mL. Infection was set to achieve a 50% of viral induced cytopathic effect measured with Cell Titer GloTM Luciferase reagent , as described in the next section.

**Neutralization Assay.**

Neutralization assays were performed in duplicate. Briefly, 60 TCID_50_ of SARS-CoV-2 were preincubated with serial dilutions of heat-inactivated plasma samples from the indicated individuals for 1 h at 37 ºC. Pre-incubated viruses were added to 60.000 Vero E6 cells per well in 96 well plates. 72 h later, viral-induced cytopathic effect was measured using the Cell Titer Glo Luciferase reagent (Promega) and the Luminoskan Plate Reader from Thermofisher. The relative light units (RLU) were normalized to untreated non-infected cells (without plasma or virus), and the ID_50_ (the reciprocal dilution inhibiting 50% of the cytopathic effect) was calculated by plotting and fitting the log of plasma dilution vs. response to a 4-parameter equation in GraphPad Prism 9.3.1, as previously described in (2, 3, 4).

1. Rodon, J., Muñoz-Basagoiti, J., Perez-Zsolt, D., Noguera-Julian, M., Paredes, R., Mateu, L., Quiñones, C., Perez, C., Erkizia, I., Blanco, I., Valencia, A., Guallar, V., Carrillo, J., Blanco, J., Segalés, J., Clotet, B., Vergara-Alert, J., Izquierdo-Useros, N., 2021. Identification of Plitidepsin as Potent Inhibitor of SARS-CoV-2-Induced Cytopathic Effect After a Drug Repurposing Screen. Front. Pharmacol. 12, 646676. <https://doi.org/10.3389/fphar.2021.646676>

2. Trinité B, Tarrés-Freixas F, Rodon J, Pradenas E, Urrea V, Marfil S, Rodríguez de la Concepción ML, Ávila-Nieto C, Aguilar-Gurrieri C, Barajas A, et al. SARS-CoV-2 infection elicits a rapid neutralizing antibody response that correlates with disease severity. *Sci Rep* (2021) 11:2608. doi:10.1038/s41598-021-81862-9

3. Trinité B, Pradenas E, Marfil S, Rovirosa C, Urrea V, Tarrés-Freixas F, Ortiz R, Rodon J, Vergara-Alert J, Segalés J, et al. Previous SARS-CoV-2 Infection Increases B.1.1.7 Cross-Neutralization by Vaccinated Individuals. *Viruses* (2021) 13:1135. doi:10.3390/v13061135

4. Pradenas E, Trinité B, Urrea V, Marfil S, Ávila-Nieto C, Rodríguez de la Concepción ML, Tarrés-Freixas F, Pérez-Yanes S, Rovirosa C, Ainsua-Enrich E, et al. Stable neutralizing antibody levels six months after mild and severe COVID-19 episode. *Med* (2021)S2666634021000350. doi:10.1016/j.medj.2021.01.005

**3-Figure.**

Total SARS-CoV-2 specific T-cell responses measured by IFN-γ ELISPOT at screening, 21 days after first vaccination (day 21), 14 days after second vaccination (day 35), vaccine group and variant of concern assessed expressed as mean spot forming cells / 106 PBMC (95% CI)

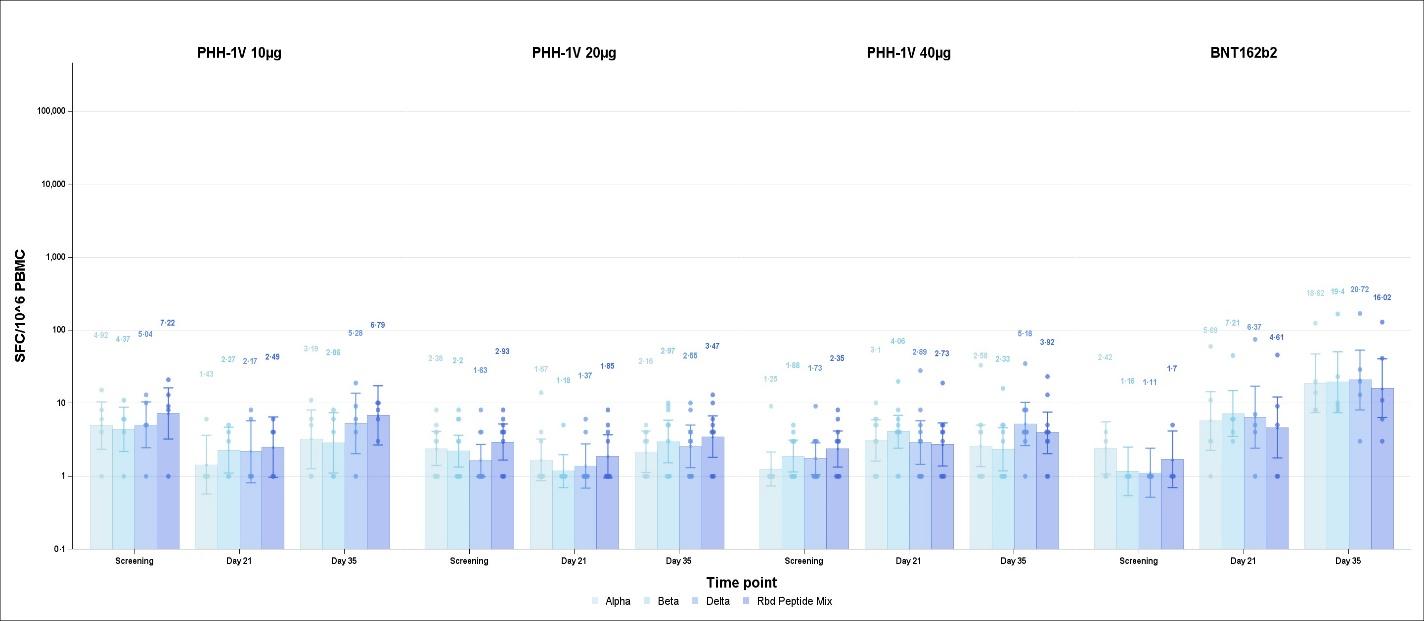

**4-HIPRA-HH-1V study group**

- Hospital Clínic Barcelona, IDIBAPS, Universitat de Barcelona, Barcelona Spain: Eva Bonfill, Omar Anagua, Faisury Caicedo, Clara Castán, Fauno Guazina, Sara Messeguer, Marta Aldea, Anna Vilella, Sandra Serrano, Lorna Leal PhD, Judit Pich, Jocelyn Nava, Karla Escobar, Joan Albert Arnaiz, Alex Soriano, José Ríos, Teresa Botta, Ignasi Esteban, Carmen Pastor, Montserrat Plana, Gemma Domenech.

- IRSICAIXA: Silvia Marfil, Carla Rovirosa, Raquel Ortiz, Daniel Perez-Zsolt, Marçal Gallemí, Edwards Pradenas, Dàlia Raïch-Regué, Benjamin Trinité, Jordana Muñoz-Basagoiti, Bonaventura Clotet, Nuria Izquierdo-Useros, Julià Blanco.

- Institut Universitari d'Investigació en Atenció Primària Jordi Gol (IDIAP Jordi Gol), Biomedical Research Institute (IdIBGi), Girona, Spain: Marina González del Río Ruth Martí-Lluch Marc Ribas-Aulinas Aida Corpes-Comes, Marga Nadal, Rafel Ramos.

- HIPRA: Luís González, Manuel Cañete, Laia Madrenas, Alexandra Moros, Irina Güell, Laura Ferrer, Antoni Prenafeta, Júlia Corominas, Carme Garriga, Antonio Barreiro, Teresa Prat, Elia Torroella.
